# Blood transcriptomic biomarkers of alcohol consumption and cardiovascular disease risk factors: the Framingham Heart Study

**DOI:** 10.1101/2022.03.08.22272112

**Authors:** Jiantao Ma, Allen Huang, Kaiyu Yan, Xianbang Sun, Roby Joehanes, Tianxiao Huan, Daniel Levy, Chunyu Liu

## Abstract

**Background:** The relations of alcohol consumption and gene expression remain to be elucidated in large study samples.

**Objectives:** To study the relationship between alcohol consumption, gene expression, and cardiovascular disease (CVD) risk factors.

**Methods:** We performed cross-sectional association analysis of whole blood derived gene expression levels with alcohol consumption in 5,531 Framingham Heart Study (FHS) participants (mean age 55 years; 54% women) by splitting the sample into a discovery sample and a replication sample (2:1 ratio) using independent pedigrees. We also examined the cross-sectional association of alcohol-associated genes with three CVD risk factors: obesity, hypertension, and diabetes. Linear mixed models or generalized estimation equations were used to quantify associations accounting for familial relationship and multiple covariates.

**Results:** We identified 25 alcohol-associated genes (false discovery rate < 0.05 in the discovery sample and Bonferroni corrected *P* < 0.05 in the replication sample). We further showed cross-sectional associations of 16 alcohol-associated genes with obesity, nine genes with hypertension, and eight genes with diabetes (all *P* < 0.002). For example, we observed decrease in expression of *PROK2* (β = -0.0018; 95%CI: -0.0021, -0.0007; P = 6.5e-5) and *PAX5* (β = -0.0014; 95%CI: -0.0021, -0.0007; P = 6.5e-5) per 1 g/day increase in alcohol consumption. Consistent with our previous observation on the inverse association of alcohol consumption with obesity and positive association of alcohol consumption with hypertension, we found that *PROK2* was positively associated with obesity (OR = 1.42; 95%CI: 1.17, 1.72; *P* = 4.5e-4) and *PAX5* was negatively associated with hypertension (OR = 0.73; 95%CI: 0.59, 0.89; *P* = 1.6e-3). We also observed that alcohol consumption was positively associated with expression of *ABCA13* (β = 0.0012; 95%CI: 0.0007, 0.0017; P = 1.3e-6) and *ABCA13* was positively associated with diabetes (OR = 2.57; 95%CI: 1.73, 3.84; *P* = 3.5e-06); this finding, however, was inconsistent with our observation on the inverse association between alcohol consumption and diabetes.

**Conclusions:** We observed that alcohol consumption was associated with whole blood expression levels of 25 genes in middle aged to older adults in the FHS. In addition, complex relationships may exist between alcohol-associated genes and CVD risk factors.

## Introduction

Excessive alcohol consumption is common and it is associated with 2.5 million deaths each year, imposing a great public health burden.[1-3] Previous studies showed that excessive alcohol consumption raises blood pressure, causes diabetes, and increases blood triglyceride levels;[4, 5] alcohol consumption therefore is an important modifiable lifestyle risk factor for cardiovascular disease (CVD). Earlier studies supported the dogma that light-to-moderate alcohol consumption is associated with reduced CVD risk; several recent studies, however, failed to show the beneficial relations of light-to-moderate consumption with reduced risk for CVD.[6-8] Multiple environmental and biological factors may be associated with such contradictory observations.

Gene expression is the process by which the genetic code – the DNA sequence – directs protein synthesis. Variations in gene expression underlie differences in cellular phenotypes, and analysis of gene expression in relation to disease phenotypes may provide insights of molecular mechanisms of diseases.[9] The use of high-throughput techniques in association studies has demonstrated the important role of perturbed gene expression in the pathogenesis of complex diseases including CVD.[10-12]. Integration of gene expression or transcriptomic data may help us to better understand the relationship between alcohol consumption and multiple diseases including CVD.[13, 14] For example, an alcoholism case-control study showed upregulation of genes involved in pathways related to inflammation, hypoxia, and stress and downregulation of genes in neurogenesis and myelination related pathways in alcoholism.[15] This evidence suggests that identifying gene expression levels associated with alcohol consumption may help better understand the effects of alcohol consumption on the risk of diseases.[15-17] Nevertheless, to the best of our knowledge, such an approach has not been systematically conducted to explore the potential effects of alcohol consumption on CVD risk in the general population.

Microarray-based gene expression profiling was performed on whole blood derived DNA collected from 5,626 Framingham Heart Study (FHS).[18, 19] In this study, we aimed to identify gene expression signatures of alcohol consumption. Our aims were to identify genome-wide gene expression changes associated with alcohol consumption and to investigate whether alcohol-associated gene expression levels were associated with CVD risk factors including obesity, hypertension, and diabetes.

## METHODS

### Study participants

The present study included participants who attended the eighth examination (n = 2,971; 2005 to 2008) of the FHS Offspring cohort and the second examination (n = 3,357; 2008 to 2011) of the FHS Third Generation cohort. [20, 21] We excluded participants who had missing values in gene expression, alcohol consumption, and any of the covariates. For the purpose of internal validation, we split our study sample into two independent groups: discovery and replication samples with a sample size ratio of 2:1 by independent pedigrees. The discovery set included 3,655 participants and the replication set included 1,876 participants. The Framingham Heart Study protocols and procedures were approved by the Institutional Review Board for Human Research at Boston University Medical Center. All participants provided written consent.

### Alcohol consumption phenotypes

At each health exam, the alcohol consumption information was collected as average drinks of beers, white and red wine, and spirits consumed in the past year using standard FHS questionnaires. The continuous phenotype, “grams of alcohol consumed per day”, was calculated using the following converters between drinks and grams: one beer (12 oz) ∼14 g ethanol, or one glass of wine (5 oz) ∼ 15 g ethanol, or one drink of spirit (1.5 oz 80 proof alcohol) ∼ 14 g ethanol. Based on the numbers of alcohol drinks consumed per week, we group study participants as nondrinkers, light to moderate drinkers (i.e., 0.1 to 14 drinks per week in women or 0.1 to 21 drinks per week in men), and heavy drinkers (i.e., ≥14 drinks per week in women or ≥21 drinks per week in men. In this study, the alcohol consumption phenotypes were selected as contemporaneous to gene expression measurements.

### Gene expression profiling

Commercial microarray analysis of gene expression in the FHS participants was conducted as described previously.[22] In brief, fasting peripheral whole blood samples were collected in PAXgene™ tubes. RNA was isolated (Asuragen, Inc.) following standard operating procedures using a KingFisher^®^ 96 robot. 50 ng RNA samples were amplified to create the cDNA library, which was then processed in the Affymetrix 7G GCS3000 scanner annotated with the Human Exon 1.0 ST Array probeset. The final gene expression profile was residualized through a linear mixed model with adjustment for 10 significant technical covariates and other factors as fixed effects as well as batch as a random effect.[22]

### CVD risk factors

At each physical and clinical examination, demographic variables and CVD risk factors were collected from all FHS participants. We analyzed three major CVD risk factors. Body mass index (BMI) was calculated as weight (kg) divided by square of height (m^2^). Obesity was defined by BMI ≥ 30 kg/m^2^. Hypertension was defined by systolic blood pressure (SBP) ≥ 140 mm Hg or diastolic blood pressure (DBP) ≥ 90 mm Hg or taking antihypertensive drugs to lower high blood pressure. Diabetes was defined by fasting blood glucose level ≥ 126 mg/dL or taking antidiabetic drugs to lower blood glucose levels.

### Statistical Analysis

We conducted a series of association and bioinformatic analyses to examine the relations between alcohol consumption, gene expression, and CVD risk factors (**Figure 1**). All association analyses were performed using R software (version 4.05).

**Figure 1.**
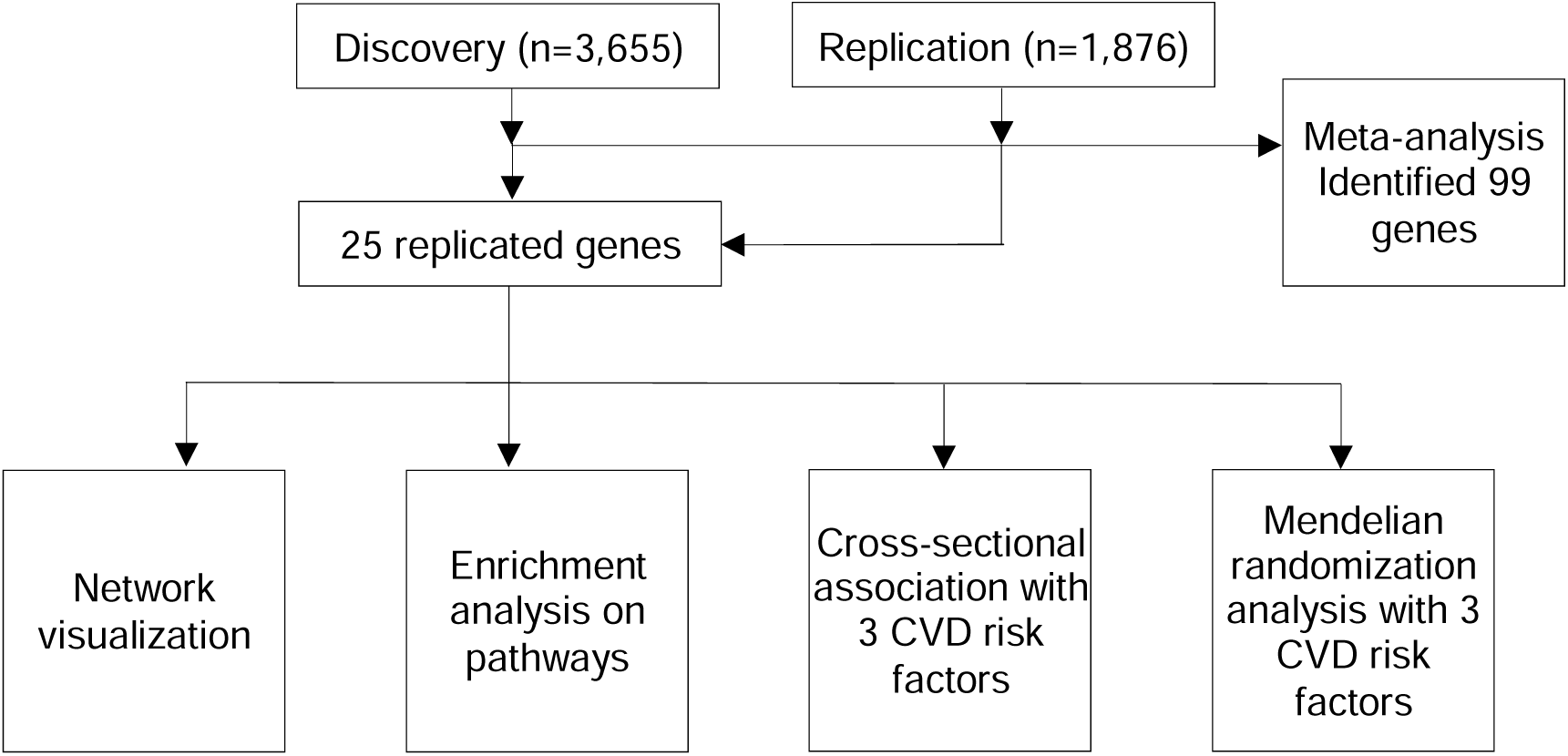
Study design flow-chart.

#### Association analysis between alcohol consumption and gene expression levels

Gene expression levels were dependent variables and alcohol consumption (grams/day) was the main independent variable in association analyses. Linear mixed effects models were used to account for family structure in FHS participants. Covariates included age, sex, BMI, current smoking status, the FHS cohort index, and blood cell counts (counts of white cell, red cell, and platelet and proportion of neutrophils, lymphocytes, monocytes, basophils, and eosinophils [22]). We considered a false discovery rate (FDR) < 0.05 for significance in the discovery sample. The selected genes from the discovery analysis were further tested in the replication sample. We used Bonferroni correct *P* < 0.05/n for significance in the replication analyses. Here n is the number of genes selected in the discovery phase. We performed an inverse variance fixed-effect meta-analysis to summarize results from the discovery and replication phases. We considered an alcohol-gene pair with Bonferroni corrected *P* <0.05/n, where n is the number of all genes tested, significant in the meta-analysis. To reduce the potential bias due to high heterogeneity, we also required that a significant alcohol-gene pair must have I^2^ <50%. Further, in the sensitivity analysis, we excluded heavy drinkers (n=304; 5.5% of overall study participants) to test if the alcohol-gene association was driven by heavy alcohol consumption.

#### Association analysis between alcohol-associated gene expression levels with CVD risk factors

We performed cross-sectional association analyses of alcohol-associated gene expression levels with three prevalent CVD risk factors, including obesity, hypertension, and diabetes. We used generalized estimating equations to quantify the associations of alcohol-associated gene expression markers as independent variables and prevalent CVD risk factors as outcome variables accounting for family structure. Covariates included age, sex, BMI (only for hypertension and diabetes), white blood cell counts and differential counts, and the FHS cohort index. Bonferroni correction was used for significance to correct for the number of genes tested for an outcome.

#### Network and enrichment analyses

We used the R *igraph* package to visualize gene clusters associated with alcohol consumption. The Kamada-Kawai layout algorithm with optimal community structure was used to demonstrate the network.[23] In this analysis, we grouped genes that had absolute pair-wise Pearson correlation coefficients ≥0.6. We used the platform of the Functional Mapping and Annotation of Genome-Wide Association Studies (FUMA GWAS) to conduct gene set enrichment analysis.[24, 25] We used the protein-coding genes as background genes. Hypergeometric tests were performed to derive *P* values for overrepresentation in gene sets obtained from databases such as MsigDB, WikiPathways, and genome-wide association studies (GWAS)-catalog. We reported significant pathways with FDR < 0.05.

#### Mendelian randomization analyses

To test the putative causal relationship between alcohol-associated genes and CVD risk factors (BMI, systolic blood pressure, diastolic blood pressure, and type 2 diabetes), we used the R *TwoSampleMR* package to perform Mendelian randomization (MR) analysis.[26] We performed the primary analysis using the inverse variance weighted (IVW) method and sensitivity analysis using the MR-Egger method. We used independent *cis*-eQTLs (i.e., genetic variants associated with gene expression in *cis* location and pair-wise linkage disequilibrium or LD r^2^ <0.1) as instrumental variables (IVs). The effect sizes and standard errors for IV-gene associations were obtained from the FHS and the effect sizes and standard errors for associations between IVs and CVD risk factors were obtained from published large GWAS.[27-29]

## Results

### Participant characteristics

The mean age of participants was 55 years and 54% were women in both the discovery and replication sets (**Table 1**). The median daily alcohol consumption was also similar between the two sets (4.9 g/day in the discovery cohort and 4.3 g/day in the validation set; Mann-Whitney-Wilcoxon Test *P* = 0.06). In addition, the means of gene expression were similar between the discovery and replication sets (Pearson correlation coefficients ≈ 0.99).

**Table 1.** Participant characteristics

| Variable | Total Sample | Discovery Set | Replication set |
| --- | --- | --- | --- |
| N | 5531 | 3655 | 1876 |
| Age | 55.4 ± 13.1 | 55.4 ± 13.1 | 55.4 ± 13.1 |
| Women | 3002 (54.3%) | 1989 (54.4%) | 1013 (54.0%) |
| Body mass index (kg/m<sup>2</sup>) | 28.18 ± 5.7 | 28.1 ± 5.5 | 28.34 ± 6.0 |
| Obesity | 1705 (30.8%) | 1122 (30.7%) | 605 (32.2%) |
| Systolic blood pressure (mm Hg) | 121.6 ± 16.8 | 121.42 ± 16.8 | 122.02 ± 16.9 |
| Diastolic blood pressure (mm Hg) | 73.94 ± 9.8 | 73.96 ± 9.7 | 73.89 ± 10.1 |
| Hypertension | 2128 (38.5%) | 1379 (37.7%) | 749 (39.3%) |
| Blood pressure medication | 1965 (35.5%) | 1280 (35.0%) | 685 (36.5%) |
| Fasting Serum glucose (mg/dL) | 100.96± 21.6 | 100.86± 21.2 | 101.15 ± 22.4 |
| Type 2 Diabetes | 493 (8.9%) | 310 (8.5%) | 183 (9.8%) |
| Diabetes medication | 881 (15.9%) | 554 (15.2%) | 327 (17.4%) |
| Current smoking | 475 (8.6%) | 330 (9.0%) | 145 (7.7%) |
| Daily alcohol consumption(g/d) | 4.3 (15.1) | 4.9 (14.9) | 4.3 (14.4) |
| Non-drinkers | 1755(31.7%) | 1132(31.0%) | 623 (33.2%) |

For continuous variables, mean (SD) is presented. For binary variables, n (%) is presented. The discovery and replication sets are independent with regard to families.

### Association analyses of alcohol consumption with gene expression levels

In the discovery set, 268 autosomal genes were associated with alcohol consumption at FDR < 0.05 (corresponding to *P* < 8e-4; **Supplemental Table 1**). In the replication set, 143 of the 268 genes were associated with alcohol consumption with *P* < 0.05 and in the same direction as that in the discovery analysis; 25 of the 143 genes were associated with alcohol consumption after Bonferroni correction (corresponding *P* < 1.9e-4; **Table 2**; **Supplemental Figure 1**). For these 25 alcohol-associated genes, alcohol consumption was positively associated with expression levels of 10 genes and negatively associated with expression levels of 15 genes (**Figure 2**). For example, for 1 g/day increase in alcohol consumption, expression levels of *CTSG* increased in both discovery and replication analyses, β = 0.0037; 95%CI: 0.0026, 0.0048; *P* = 4.0e-11 and β = 0.0033; 95%CI: 0.0019, 0.0047; *P* = 4.7e-6, respectively; expression levels of *PROK2* decreased in both discovery and replication analyses, β = -0.0018; 95%CI: -0.0025, -0.0012; *P* = 1.1e-07 and β = -0.0020; 95%CI: -0.0029, -0.0011; *P* = 2.1e-05, respectively. In sensitivity analysis after heavy drinkers were excluded, the regression coefficients were comparable to those from analyses in all study participants (**Supplemental Figure 2**).

**Table 2.**
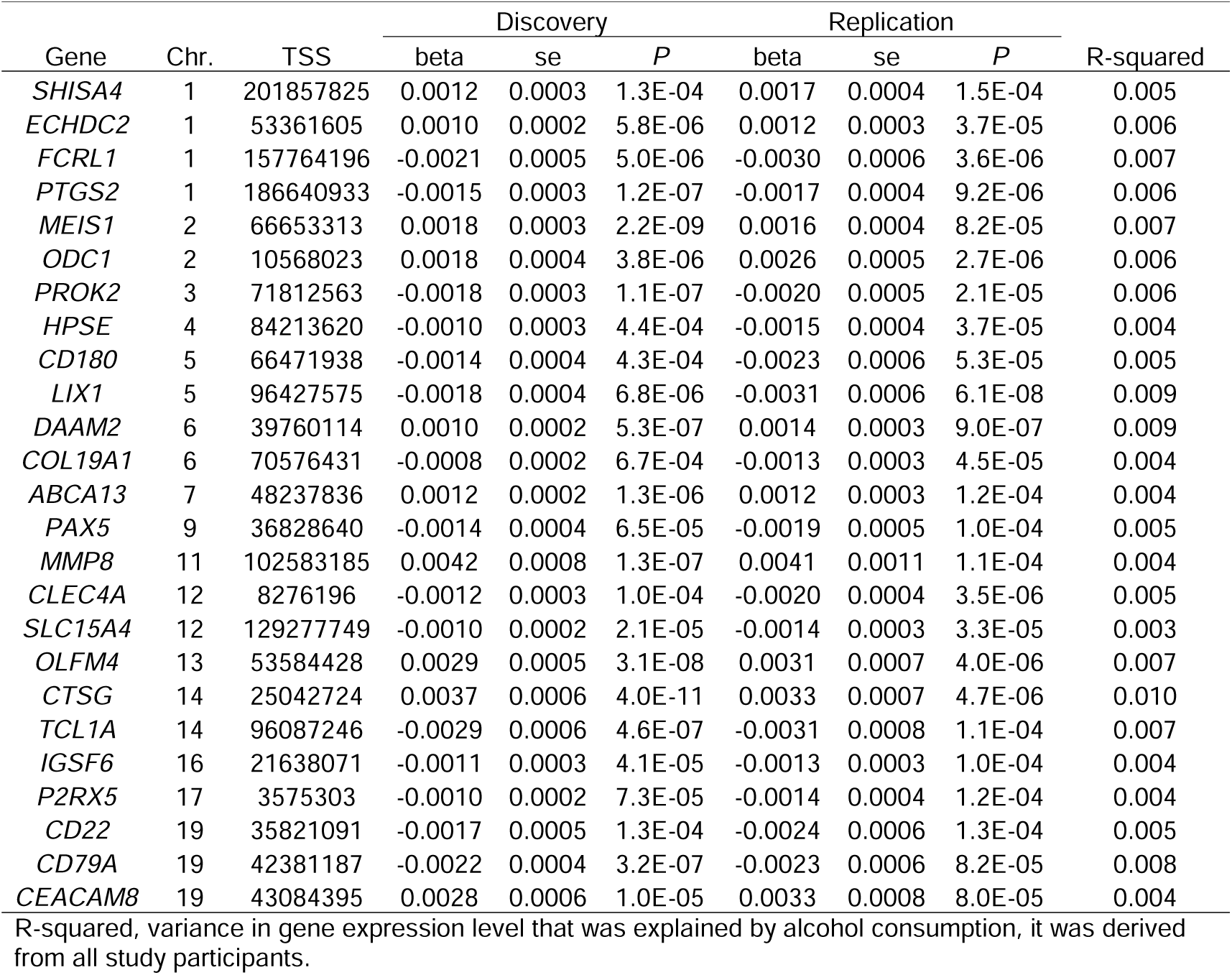
Cross-sectional associations of alcohol consumption with 25 genes

| Gene | Chr. | TSS | Discovery |  |  | Replication |  |  | R-squared |
| --- | --- | --- | --- | --- | --- | --- | --- | --- | --- |
|  |  |  | beta | se | P | beta | se | P |  |
| *SHISA4* | 1 | 201857825 | 0.0012 | 0.0003 | 1.3E-04 | 0.0017 | 0.0004 | 1.5E-04 | 0.005 |
| *ECHDC2* | 1 | 53361605 | 0.0010 | 0.0002 | 5.8E-06 | 0.0012 | 0.0003 | 3.7E-05 | 0.006 |
| *FCRL1* | 1 | 157764196 | -0.0021 | 0.0005 | 5.0E-06 | -0.0030 | 0.0006 | 3.6E-06 | 0.007 |
| *PTGS2* | 1 | 186640933 | -0.0015 | 0.0003 | 1.2E-07 | -0.0017 | 0.0004 | 9.2E-06 | 0.006 |
| *MEIS1* | 2 | 66653313 | 0.0018 | 0.0003 | 2.2E-09 | 0.0016 | 0.0004 | 8.2E-05 | 0.007 |
| *ODC1* | 2 | 10568023 | 0.0018 | 0.0004 | 3.8E-06 | 0.0026 | 0.0005 | 2.7E-06 | 0.006 |
| *PROK2* | 3 | 71812563 | -0.0018 | 0.0003 | 1.1E-07 | -0.0020 | 0.0005 | 2.1E-05 | 0.006 |
| *HPSE* | 4 | 84213620 | -0.0010 | 0.0003 | 4.4E-04 | -0.0015 | 0.0004 | 3.7E-05 | 0.004 |
| *CD180* | 5 | 66471938 | -0.0014 | 0.0004 | 4.3E-04 | -0.0023 | 0.0006 | 5.3E-05 | 0.005 |
| *LIX1* | 5 | 96427575 | -0.0018 | 0.0004 | 6.8E-06 | -0.0031 | 0.0006 | 6.1E-08 | 0.009 |
| *DAAM2* | 6 | 39760114 | 0.0010 | 0.0002 | 5.3E-07 | 0.0014 | 0.0003 | 9.0E-07 | 0.009 |
| *COL19A1* | 6 | 70576431 | -0.0008 | 0.0002 | 6.7E-04 | -0.0013 | 0.0003 | 4.5E-05 | 0.004 |
| *ABCA13* | 7 | 48237836 | 0.0012 | 0.0002 | 1.3E-06 | 0.0012 | 0.0003 | 1.2E-04 | 0.004 |
| *PAX5* | 9 | 36828640 | -0.0014 | 0.0004 | 6.5E-05 | -0.0019 | 0.0005 | 1.0E-04 | 0.005 |
| *MMP8* | 11 | 102583185 | 0.0042 | 0.0008 | 1.3E-07 | 0.0041 | 0.0011 | 1.1E-04 | 0.004 |
| *CLEC4A* | 12 | 8276196 | -0.0012 | 0.0003 | 1.0E-04 | -0.0020 | 0.0004 | 3.5E-06 | 0.005 |
| *SLC15A4* | 12 | 129277749 | -0.0010 | 0.0002 | 2.1E-05 | -0.0014 | 0.0003 | 3.3E-05 | 0.003 |
| *OLFM4* | 13 | 53584428 | 0.0029 | 0.0005 | 3.1E-08 | 0.0031 | 0.0007 | 4.0E-06 | 0.007 |
| *CTSG* | 14 | 25042724 | 0.0037 | 0.0006 | 4.0E-11 | 0.0033 | 0.0007 | 4.7E-06 | 0.010 |
| *TCL1A* | 14 | 96087246 | -0.0029 | 0.0006 | 4.6E-07 | -0.0031 | 0.0008 | 1.1E-04 | 0.007 |
| *IGSF6* | 16 | 21638071 | -0.0011 | 0.0003 | 4.1E-05 | -0.0013 | 0.0003 | 1.0E-04 | 0.004 |
| *P2RX5* | 17 | 3575303 | -0.0010 | 0.0002 | 7.3E-05 | -0.0014 | 0.0004 | 1.2E-04 | 0.004 |
| *CD22* | 19 | 35821091 | -0.0017 | 0.0005 | 1.3E-04 | -0.0024 | 0.0006 | 1.3E-04 | 0.005 |
| *CD79A* | 19 | 42381187 | -0.0022 | 0.0004 | 3.2E-07 | -0.0023 | 0.0006 | 8.2E-05 | 0.008 |
| *CEACAM8* | 19 | 43084395 | 0.0028 | 0.0006 | 1.0E-05 | 0.0033 | 0.0008 | 8.0E-05 | 0.004 |

R-squared, variance in gene expression level that was explained by alcohol consumption, it was derived from all study participants.

**Figure 2.**
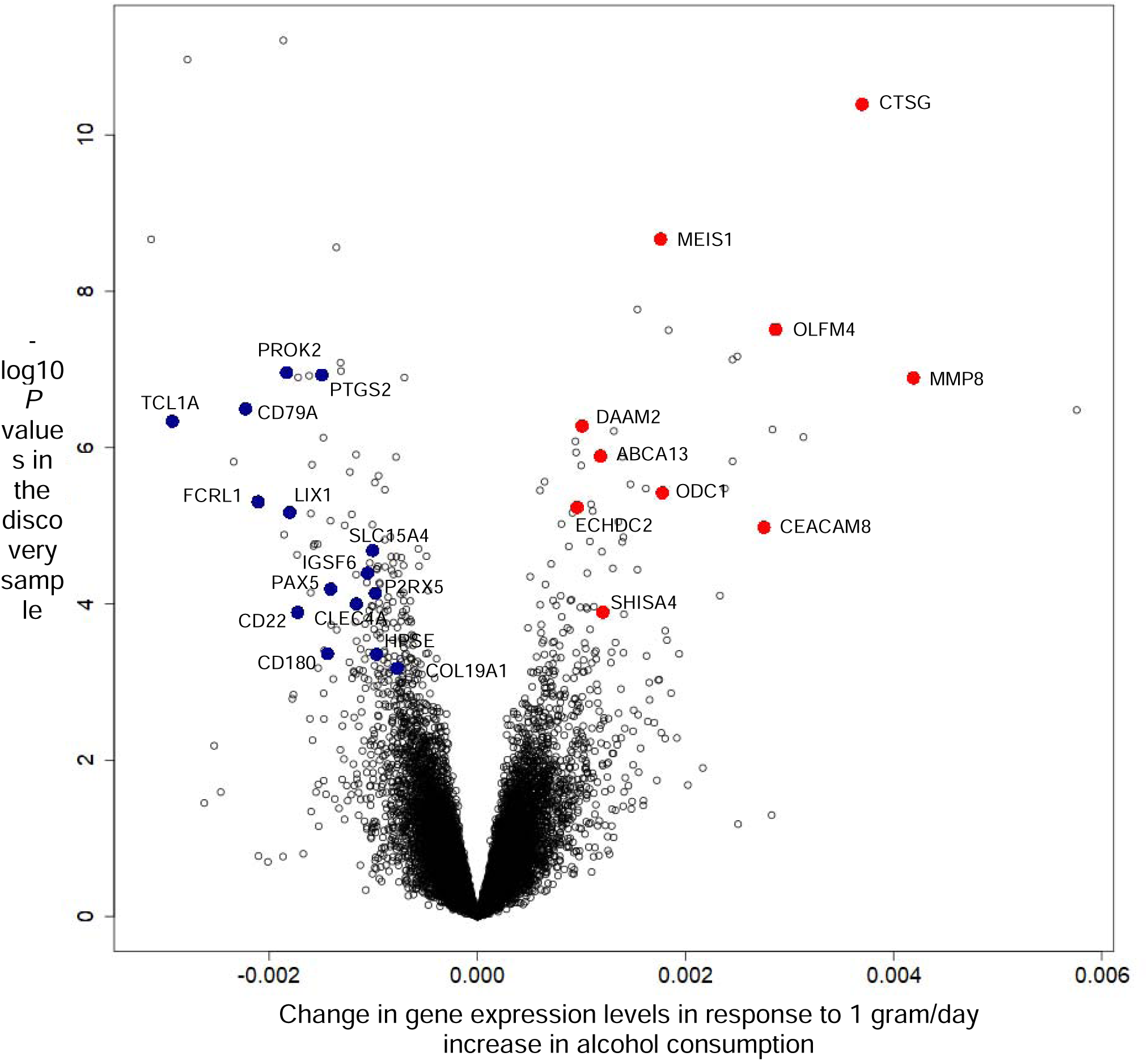
Volcano plot for the association analysis of alcohol consumption and whole blood derived genes in the discovery sample.

Pair-wise correlations of expression for the 25 alcohol-associated genes (n = 300 pairs) were low to moderate, 84.7% of gene pairs (n = 254) had Pearson |*r*| < 0.3 (**Supplemental Figure 3**). Using modularity optimization to visualize network of the alcohol-associated genes, we found two alcohol-associated gene clusters after removing the low correlation (|*r*| < 0.6) edges (**Supplemental Figure 4**). One cluster included five genes (*CTSG, MMP8, ABCA13, CEACAM8*, and *OLFM4*) and the other cluster included six genes (*TCL1A, PAX5, CD79A, CD22, FCRL1*, and *COL19A1*). Enrichment analysis showed 66 significant pathways at FDR < 0.05 (**Supplemental Table 2**). For example, the 25 alcohol-associated genes were involved in six gene ontology (GO) biological processes such as cell activation in immune response (GO:0002263; FDR = 0.006; corresponding *P* = 1.8e-6), myeloid leukocyte mediated immunity (GO:0002444; FDR = 0.02; corresponding *P* = 4.3e-6) , and immune effector process (GO:0002252; FDR = 0.01; corresponding *P* = 7.6e-6). Enrichment analysis also showed that the two clusters of alcohol-associated genes identified in network visualization were likely involved in inflammation related pathways.

Using fixed-effect meta-analysis to summarize the associations of gene expression and alcohol consumption in discovery and replication samples, we founded that the levels of 99 autosomal genes were associated with alcohol consumption after Bonferroni correction (corresponding *P* < 2.8e-6; **Supplemental Table 3**). The 99 genes included 23 (92%) of the 25 alcohol-associated genes identified using the discovery-replication approach. In meta-analyses, the heterogeneity of most of the 99 associations was low between the discovery and replication cohorts: 93 alcohol-gene pairs with I^2^ ≤ 0.3 and the other 6 pairs with I^2^ ranging from 0.31 to 0.49 . We conducted enrichment analysis and found 199 significant pathways at FDR < 0.05 (**Supplemental Table 4**). In this analysis with the 99 genes, we also observed enrichment of additional inflammation related pathways such as GSE4984_UNTREATED_VS_GALECTIN1_TREATED_DC_DN (genes down-regulated in monocyte-derived dendritic cells: control versus treated with *LGALS1*; *P* = 5.2e-11; FDR = 2.0e-7)

### Alcohol-associated genes and CVD risk factors

We performed cross-sectional association analyses of the 25 alcohol-associated genes identified in the discovery and replication analyses (*P* < 0.002) with the three CVD risk factors. We found that 16 genes were associated with obesity, nine genes with hypertension, and eight genes with diabetes (**Table 3**). Our previous study in the FHS has shown that alcohol consumption was inversely associated with obesity and diabetes while positively associated with hypertension.[30] If alcohol consumption was positively associated with expression levels of a gene, this gene was therefore expected to be positively associated with hypertension or negatively associated with obesity or diabetes. The directions of the nine associations between alcohol-associated genes and hypertension were all consistent with the *a priori* expectation. For example, in the discovery analysis, we observed that that 1 g/day higher alcohol consumption was associated with 0.0014 units lower expression levels of *PAX5* (95%CI: -0.0021, -0.0007; *P* = 6.5e-5). Consistent with this finding, we found that one-unit lower expression level of *PAX5* was associated with 37% higher odds of hypertension (odds ratio or OR = 1.38, 95%CI: 1.13, 1.68; *P* = 1.6e-3). About half of the gene-obesity pairs (nine of 16 alcohol-associated genes) showed consistent direction while none of the gene-diabetes pairs were consistent with the expectation. For example, in the discovery analysis, we observed that alcohol consumption was negatively associated with the expression level of *PROK2* (β = -0.0018; 95%CI: -0.0025, - 0.0012; *P* = 1.1e-7) and positively associated with expression of *ABCA13* (β = 0.0012; 95%CI: 0.0007, 0.0017; P = 1.3e-6). Consistently with the association of alcohol consumption with *PROK2*, we found that *PROK2* was positively associated with obesity (OR = 1.42; 95%CI: 1.17, 1.72; *P* = 4.5e-4). Whereas we found that *ABCA13* was positively associated with diabetes (OR = 2.57; 95%CI: 1.73, 3.84; *P* = 3.5e-06), and this is inconsistent with the observed positive association between alcohol consumption and *ABCA13*.

**Table 3.** Cross-sectional analysis of the 25 alcohol associated genes with CVD risk factors

| Gene | Obesity |  |  | Hypertension |  |  | Diabetes |  |  |
| --- | --- | --- | --- | --- | --- | --- | --- | --- | --- |
|  | beta | se | P | beta | se | P | beta | se | P |
| *SHISA4* | 0.98 | 1.11 | 8.6E-01 | 1.50 | 1.12 | 5.9E-04 * | 1.04 | 1.20 | 8.3E-01 |
| *ECHDC2* | 1.95 | 1.17 | 2.4E-05 * | 1.56 | 1.19 | 1.2E-02 | 1.19 | 1.30 | 5.1E-01 |
| *FCRL1* | 1.26 | 1.07 | 1.2E-03 * | 0.88 | 1.08 | 1.1E-01 | 0.92 | 1.12 | 4.9E-01 |
| *PTGS2* | 0.67 | 1.12 | 5.2E-04 * | 0.53 | 1.14 | 4.4E-07 * | 0.41 | 1.21 | 2.6E-06 * |
| *MEIS1* | 1.18 | 1.12 | 1.4E-01 | 1.57 | 1.13 | 2.8E-04 * | 0.96 | 1.21 | 8.3E-01 |
| *ODC1* | 2.04 | 1.09 | 2.1E-16 * | 1.74 | 1.10 | 5.9E-09 * | 2.46 | 1.17 | 7.4E-09 * |
| *PROK2* | 1.42 | 1.10 | 4.5E-04 * | 0.92 | 1.11 | 4.2E-01 | 1.49 | 1.21 | 3.4E-02 |
| *HPSE* | 2.14 | 1.14 | 2.7E-09 * | 0.95 | 1.14 | 6.9E-01 | 1.05 | 1.22 | 8.0E-01 |
| *CD180* | 1.92 | 1.09 | 3.2E-15 * | 1.07 | 1.09 | 4.3E-01 | 1.24 | 1.14 | 1.2E-01 |
| *LIX1* | 1.71 | 1.08 | 1.8E-11 * | 0.97 | 1.09 | 7.5E-01 | 1.25 | 1.15 | 1.1E-01 |
| *DAAM2* | 0.24 | 1.21 | 2.7E-13 * | 0.96 | 1.19 | 8.4E-01 | 0.33 | 1.36 | 3.1E-04 * |
| *COL19A1* | 1.90 | 1.15 | 7.0E-06 * | 0.78 | 1.18 | 1.2E-01 | 0.69 | 1.28 | 1.3E-01 |
| *ABCA13* | 2.29 | 1.15 | 4.8E-09 * | 1.40 | 1.17 | 3.0E-02 | 2.57 | 1.23 | 3.5E-06 * |
| *PAX5* | 0.96 | 1.10 | 6.5E-01 | 0.73 | 1.11 | 1.6E-03 * | 0.74 | 1.17 | 6.2E-02 |
| *MMP8* | 1.61 | 1.04 | 5.2E-28 * | 1.27 | 1.05 | 2.6E-07 * | 1.51 | 1.06 | 1.9E-11 * |
| *CLEC4A* | 1.17 | 1.11 | 1.5E-01 | 0.85 | 1.13 | 1.8E-01 | 1.05 | 1.20 | 7.8E-01 |
| *SLC15A4* | 0.73 | 1.15 | 2.0E-02 | 0.58 | 1.16 | 2.5E-04 * | 0.51 | 1.27 | 5.5E-03 |
| *OLFM4* | 1.50 | 1.07 | 3.3E-10 * | 1.17 | 1.07 | 2.8E-02 | 1.55 | 1.10 | 7.7E-06 * |
| *CTSG* | 1.18 | 1.06 | 4.7E-03 | 1.17 | 1.07 | 2.0E-02 | 1.37 | 1.10 | 5.8E-04 * |
| *TCL1A* | 1.17 | 1.06 | 7.1E-03 | 0.87 | 1.06 | 1.9E-02 | 0.81 | 1.10 | 2.8E-02 |
| *IGSF6* | 1.67 | 1.14 | 7.2E-05 * | 1.08 | 1.15 | 6.0E-01 | 1.02 | 1.28 | 9.4E-01 |
| *P2RX5* | 0.98 | 1.14 | 8.9E-01 | 0.72 | 1.16 | 2.8E-02 | 1.42 | 1.27 | 1.4E-01 |
| *CD22* | 1.50 | 1.08 | 4.3E-08 * | 0.96 | 1.09 | 6.2E-01 | 1.11 | 1.13 | 4.0E-01 |
| *CD79A* | 0.95 | 1.08 | 5.4E-01 | 0.75 | 1.09 | 6.1E-04 * | 0.86 | 1.14 | 2.6E-01 |
| *CEACAM8* | 1.59 | 1.06 | 1.6E-17 * | 1.23 | 1.06 | 3.7E-04 * | 1.54 | 1.09 | 1.4E-07 * |

\*: Genes are significantly associated with CVD risk factors after Bonferroni correction

We performed MR analyses to test the putative causal relationship between alcohol-associated genes and CVD risk factors. We found six significant gene-CVD risk factors pairs at *P* <0.002 (**Table 4**). For five of the six gene-CVD risk factor pairs, the direction was consistent with the *a priori* expectation. For example, alcohol consumption was associated with lower expression levels of *HPSE* and higher expression levels of *CTSG* (**Supplemental Figure 5**). In MR analyses, the genetically predicted *HPSE* expression level was positively associated with BMI (β = 0.003, 95%CI: 0.002, 0.005, *P* = 9.3e-5); the genetically predicted *CTSG* expression level was positively associated with systolic blood pressure (β = 0.11; 95%CI: 0.04, 0.18, *P* = 1.99e-3).

**Table 4.** Mendelian randomization analysis results for the 25 alcohol-associated genes with CVD risk factors

| Gene | CVD risk factors | Number of SNPs | beta | se | P for IVW | P for pleiotropy | P for heterogeneity |
| --- | --- | --- | --- | --- | --- | --- | --- |
| *CD180* | DBP | 2 | -0.0215 | 0.0075 | 4.1E-03 |  | 0.67 |
| *CD180* | SBP | 2 | -0.0369 | 0.0130 | 4.6E-03 |  | 0.79 |
| *CD22* | T2D | 1 | -0.0052 | 0.0024 | 3.4E-02 |  |  |
| *CD79A* | BMI | 1 | 0.0009 | 0.0004 | 3.4E-02 |  |  |
| *CTSG* | SBP | 2 | 0.1103 | 0.0357 | 2.0E-03 * |  | 0.51 |
| *ECHDC2* | T2D | 6 | 0.0060 | 0.0026 | 2.4E-02 | 0.73 | 0.95 |
| *HPSE* | BMI | 2 | 0.0032 | 0.0008 | 9.3E-05 * |  |  |
| *P2RX5* | SBP | 3 | 0.0244 | 0.0099 | 1.4E-02 | 0.55 | 0.37 |
| *P2RX5* | T2D | 1 | -0.0178 | 0.0053 | 8.0E-04 * |  |  |
| *PAX5* | DBP | 1 | 0.0569 | 0.0249 | 2.2E-02 |  |  |
| *PAX5* | SBP | 1 | 0.1144 | 0.0431 | 8.0E-03 |  |  |
| *PAX5* | BMI | 1 | 0.0155 | 0.0023 | 2.1E-11 * |  |  |
| *PAX5* | T2D | 1 | 0.0314 | 0.0105 | 2.8E-03 |  |  |
| *PROK2* | DBP | 9 | -0.0046 | 0.0011 | 2.1E-05 * | 0.71 | 0.25 |
| *PROK2* | SBP | 9 | -0.0068 | 0.0020 | 8.0E-04 * | 0.77 | 0.18 |

\*: Significant after Bonferroni correction

## Discussion

In this analysis of middle to older aged participants in the community-based FHS, we observed significant cross-sectional associations of alcohol drinking with whole blood derived expression levels of 25 autosomal genes. We then tested the association of expression levels of these genes with CVD risk factors and found that 22 of the 25 genes were associated with at least one of the three tested CVD risk factors. The directions of alcohol-associated genes with hypertension were consistent with our expectation. Whereas the directions for obesity and diabetes, particularly diabetes, were not always consistent with our expectation, suggesting complex relations between alcohol consumption and these two CVD risk factors. Taken together, this study provides novel evidence indicating that alcohol intake is linked to CVD risk factors through changes in gene expression levels in whole blood.

Several studies investigated transcriptome-wide gene expression with long-term alcohol dependence or alcoholism.[31-34] Many of these studies examined gene expression levels in postmortem tissues such as brain and liver. Some of these previous studies used microarray technologies,[33, 34] and others used the RNA sequencing technology to measure gene expression.[31, 32] These previous studies provide insights with respect to the transcriptional landscape associated with long-term heavy alcohol consumption. For example, Farris et al. examined 23 individuals with alcohol use disorders and 23 sex- and age-matched healthy controls[31] and identified differentially expressed gene networks and showed that genes in these networks may be involved in pathways related to oxidation reduction, mitochondrion, and fatty acid metabolism. However, as with other transcriptome-wide studies with small sample sizes, this study essentially found no significant individual genes associated with alcohol intake.[31, 32, 34] In addition, data generated from these previous studies may not be easily generalized to individuals who drink moderate amount of alcohol. Compared to these previous studies, our study has a much larger sample size, which enable us to identify individual genes associated with alcohol consumption in a general population where the majority of the participants were nondrinkers or light to moderate drinkers.

Given the cross-sectional and observational nature of this study, we can only speculate regarding the potential mechanisms that may explain the observed findings. We found that alcohol-associated genes were enriched in inflammation-related processes. This is in line with the previous findings that excessive alcohol consumption may induce an adverse immune response and lead to the related clinical outcomes.[35, 36] In the MR analysis, we found that five gene-CVD risk factor pairs were consistent with our *a priori* expectation. Among these genes, *CTSG* encodes a serine protease (cathepsin G), which is one an effector of inflammation.[37] The expression of *CTSG* mainly occurs in polymorphonuclear neutrophils, where cathepsin G stimulates the release of cytokines and chemokines that are responsible for activation of immune response.[38] The activity of cathepsin G has been related to diseases associated with chronic inflammation such as atherosclerosis and chronic obstructive pulmonary disease.[39, 40] Expression of *HPSE* has also been linked to immune response. Another alcohol related gene, *HPSE*, encodes heparinase, which is the primary enzyme that is responsible to the degradation of heparan sulfate, a major component of basement membrane and extracellular matrix, and the release of bioactive molecules in humans.[41] The enzymatic activity of heparinase plays critical roles in a variety of pathological processes such as inflammation and atherosclerosis.[42, 43] In addition, *PAX5* and *PROK2* are genes involved in immune responses. For example, using mice models, one study showed that mutation of *Pax5* stimulated production of proinflammatory cytokine interleukin-6,[44] and another study showed *Prok2* promoted production of proinflammatory interleukin-1β and tumor necrosis factor but decreased production of anti-inflammatory interleukin-10.[45] The pathogenic activities associated with these genes seem consistent with our observation. Nonetheless, the causal relationship between alcohol consumption and expression of these genes and whether and the extent to which expression levels of these genes mediates the association between alcohol consumption and CVD risk factors warrants future studies.

Given the adverse effects of heavy alcohol drinking, Dietary Guideline for American recommends that one should not begin to drink alcohol or intentionally continue to drink as a means of improving health.[46] Moderate alcohol consumption, however, is a component of several healthy dietary patterns.[47, 48] The present study and recent studies showed that the relationship between alcohol drinking and CVD risk is complex and occasionally paradoxical with regard to the relationships of alcohol consumption, molecular pathways, and CVD risk factors.[6-8, 30] These pieces of evidence suggest that in order to establish more appropriate recommendations for alcohol consumptions, additional studies are needed to further characterize the molecular and clinical impacts of alcohol consumption on CVD risk.

To the best of our knowledge, the present study is the largest population-based study to explore the association between habitual alcohol consumption and concurrent gene expression levels in whole blood. The strengths of the present study also include the utilization of comprehensive lifestyle and clinical data in the well-characterized FHS. Several limitations warrant discussion. We examined cross-sectional association between alcohol consumption and gene expression levels, which limits our ability to infer causality of alcohol-gene expression relations. In addition, due to the lack of longitudinal data on gene expression measurements, the long-term effect of alcohol drinking on these genes and long-term biological function of these genes in humans need to be illustrated in the future. Also, gene expression levels in other tissues such as liver may be more relevant to alcohol related health outcomes and molecular consequences; however, it is not feasible to obtain gene expression levels in such tissues in population studies like FHS. In the present study, alcohol intake explains a small fraction of interindividual variation (0.3% to 0.99%) of the overall gene expression levels, which indicates that other factors may play important roles in changing gene expression levels. Previous studies reported that a number of genetic variants (i.e., eQTLs) are strongly associated with gene expression levels.[19, 49] Physiological conditions (e.g., disease status) are also associated with gene expression levels.[10] Future studies are therefore needed to examine the potential synergistic effects of alcohol consumption with other lifestyle, clinical, and genetic factors on these genes. In the present analysis, we removed heavy drinkers and found that the effect sizes of alcohol-gene associations largely remained. Nonetheless, we are conducting study in a larger and ethnicity diverse study samples to explore if a nonlinear relationship may exist between alcohol consumption and gene expression.

In conclusion, we demonstrated that alcohol consumption was associated with concurrent blood expression levels of 25 autosomal genes. We also showed that several of these alcohol-associated genes were associated with prevalent CVD risk factors. Our findings provide novel evidence to support that integrating gene expression levels may help unravel the complex relationship between alcohol consumption and CVD risk. Additional studies are warranted to replicate our findings and to examine the causal relations between alcohol consumption, particularly moderate alcohol consumption, and gene expression levels.

## Data Availability

All data produced are available online at dbGaP (Study Accession: phs000007.v32.p13)

## Reference

1. Centers for Disease C, Prevention: Alcohol-attributable deaths and years of potential life lost--United States, 2001. MMWR Morb Mortal Wkly Rep 2004, 53(37):866–870.

2. Flensborg-Madsen T, Knop J, Mortensen EL, Becker U, Gronbaek M: Amount of alcohol consumption and risk of developing alcoholism in men and women. Alcohol Alcohol 2007, 42(5):442–447.

3. Rehm J, Mathers C, Popova S, Thavorncharoensap M, Teerawattananon Y, Patra J: Global burden of disease and injury and economic cost attributable to alcohol use and alcohol-use disorders. Lancet 2009, 373(9682):2223–2233.

4. Emanuele NV, Swade TF, Emanuele MA: Consequences of alcohol use in diabetics. Alcohol Health Res World 1998, 22(3):211–219.

5. Chait A, Mancini M, February AW, Lewis B: Clinical and metabolic study of alcoholic hyperlipidaemia. Lancet 1972, 2(7767):62–64.

6. Collaborators GBDA: Alcohol use and burden for 195 countries and territories, 1990-2016: a systematic analysis for the Global Burden of Disease Study 2016. Lancet 2018, 392(10152):1015–1035.

7. Chikritzhs TN, Naimi TS, Stockwell TR, Liang W: Mendelian randomisation meta-analysis sheds doubt on protective associations between ‘moderate’ alcohol consumption and coronary heart disease. Evid Based Med 2015, 20(1):38.

8. Stockwell T, Zhao J, Panwar S, Roemer A, Naimi T, Chikritzhs T: Do “Moderate” Drinkers Have Reduced Mortality Risk? A Systematic Review and Meta-Analysis of Alcohol Consumption and All-Cause Mortality. J Stud Alcohol Drugs 2016, 77(2):185–198.

9. Cheung VG, Spielman RS: Genetics of human gene expression: mapping DNA variants that influence gene expression. Nat Rev Genet 2009, 10(9):595–604.

10. Huan T, Esko T, Peters MJ, Pilling LC, Schramm K, Schurmann C, Chen BH, Liu C, Joehanes R, Johnson AD et al: A meta-analysis of gene expression signatures of blood pressure and hypertension. PLoS Genet 2015, 11(3):e1005035.

11. Yao C, Chen BH, Joehanes R, Otlu B, Zhang X, Liu C, Huan T, Tastan O, Cupples LA, Meigs JB et al: Integromic analysis of genetic variation and gene expression identifies networks for cardiovascular disease phenotypes. Circulation 2015, 131(6):536–549.

12. Benton MC, Lea RA, Macartney-Coxson D, Carless MA, Goring HH, Bellis C, Hanna M, Eccles D, Chambers GK, Curran JE et al: Mapping eQTLs in the Norfolk Island genetic isolate identifies candidate genes for CVD risk traits. Am J Hum Genet 2013, 93(6):1087–1099.

13. Wellcome Trust Case Control C: Genome-wide association study of 14,000 cases of seven common diseases and 3,000 shared controls. Nature 2007, 447(7145):661–678.

14. Subramanian A, Tamayo P, Mootha VK, Mukherjee S, Ebert BL, Gillette MA, Paulovich A, Pomeroy SL, Golub TR, Lander ES et al: Gene set enrichment analysis: a knowledge-based approach for interpreting genome-wide expression profiles. Proc Natl Acad Sci U S A 2005, 102(43):15545–15550.

15. McClintick JN, Xuei X, Tischfield JA, Goate A, Foroud T, Wetherill L, Ehringer MA, Edenberg HJ: Stress-response pathways are altered in the hippocampus of chronic alcoholics. Alcohol 2013, 47(7):505–515.

16. Marballi K, Genabai NK, Blednov YA, Harris RA, Ponomarev I: Alcohol consumption induces global gene expression changes in VTA dopaminergic neurons. Genes Brain Behav 2016, 15(3):318–326.

17. Li J, Bardag-Gorce F, Oliva J, Dedes J, French BA, French SW: Gene expression modifications in the liver caused by binge drinking and S-adenosylmethionine feeding. The role of epigenetic changes. Genes Nutr 2010, 5(2):169–179.

18. Huan T, Zhang B, Wang Z, Joehanes R, Zhu J, Johnson AD, Ying S, Munson PJ, Raghavachari N, Wang R et al: A systems biology framework identifies molecular underpinnings of coronary heart disease. Arterioscler Thromb Vasc Biol 2013, 33(6):1427–1434.

19. Joehanes R, Zhang X, Huan T, Yao C, Ying SX, Nguyen QT, Demirkale CY, Feolo ML, Sharopova NR, Sturcke A et al: Integrated genome-wide analysis of expression quantitative trait loci aids interpretation of genomic association studies. Genome Biol 2017, 18(1):16.

20. Feinleib M, Kannel WB, Garrison RJ, McNamara PM, Castelli WP: The Framingham Offspring Study. Design and preliminary data. Prev Med 1975, 4(4):518–525.

21. Splansky GL, Corey D, Yang Q, Atwood LD, Cupples LA, Benjamin EJ, D’Agostino RB, Sr., Fox CS, Larson MG, Murabito JM et al: The Third Generation Cohort of the National Heart, Lung, and Blood Institute’s Framingham Heart Study: design, recruitment, and initial examination. Am J Epidemiol 2007, 165(11):1328–1335.

22. Joehanes R, Ying S, Huan T, Johnson AD, Raghavachari N, Wang R, Liu P, Woodhouse KA, Sen SK, Tanriverdi K et al: Gene expression signatures of coronary heart disease. Arterioscler Thromb Vasc Biol 2013, 33(6):1418–1426.

23. Brandes U, Delling D, Gaertler M, Gorke R, Hoefer M, Nikoloski Z, Wagner D: On Modularity Clustering, in IEEE Transactions on Knowledge and Data Engineering. 2008, 20(2):172–188.

24. Watanabe K, Umicevic Mirkov M, de Leeuw CA, van den Heuvel MP, Posthuma D: Genetic mapping of cell type specificity for complex traits. Nat Commun 2019, 10(1):3222.

25. Watanabe K, Taskesen E, van Bochoven A, Posthuma D: Functional mapping and annotation of genetic associations with FUMA. Nat Commun 2017, 8(1):1826.

26. Hemani G, Zheng J, Elsworth B, Wade KH, Haberland V, Baird D, Laurin C, Burgess S, Bowden J, Langdon R et al: The MR-Base platform supports systematic causal inference across the human phenome. Elife 2018, 7.

27. Evangelou E, Warren HR, Mosen-Ansorena D, Mifsud B, Pazoki R, Gao H, Ntritsos G, Dimou N, Cabrera CP, Karaman I et al: Genetic analysis of over 1 million people identifies 535 new loci associated with blood pressure traits. Nat Genet 2018, 50(10):1412–1425.

28. Yengo L, Sidorenko J, Kemper KE, Zheng Z, Wood AR, Weedon MN, Frayling TM, Hirschhorn J, Yang J, Visscher PM et al: Meta-analysis of genome-wide association studies for height and body mass index in approximately 700000 individuals of European ancestry. Hum Mol Genet 2018, 27(20):3641–3649.

29. Xue A, Wu Y, Zhu Z, Zhang F, Kemper KE, Zheng Z, Yengo L, Lloyd-Jones LR, Sidorenko J, Wu Y et al: Genome-wide association analyses identify 143 risk variants and putative regulatory mechanisms for type 2 diabetes. Nat Commun 2018, 9(1):2941.

30. Sun X, Ho JE, Gao H, Evangelou E, Yao C, Huan T, Hwang SJ, Courchesne P, Larson MG, Levy D et al: Associations of Alcohol Consumption with Cardiovascular Disease-Related Proteomic Biomarkers: The Framingham Heart Study. J Nutr 2021, 151(9):2574–2582.

31. Farris SP, Arasappan D, Hunicke-Smith S, Harris RA, Mayfield RD: Transcriptome organization for chronic alcohol abuse in human brain. Mol Psychiatry 2015, 20(11):1438–1447.

32. Slattery ML, Pellatt DF, Mullany LE, Wolff RK: Differential Gene Expression in Colon Tissue Associated With Diet, Lifestyle, and Related Oxidative Stress. PLoS One 2015, 10(7):e0134406.

33. Zhang H, Wang F, Xu H, Liu Y, Liu J, Zhao H, Gelernter J: Differentially co-expressed genes in postmortem prefrontal cortex of individuals with alcohol use disorders: influence on alcohol metabolism-related pathways. Hum Genet 2014, 133(11):1383–1394.

34. Murano T, Koshimizu H, Hagihara H, Miyakawa T: Transcriptomic immaturity of the hippocampus and prefrontal cortex in patients with alcoholism. Sci Rep 2017, 7:44531.

35. Szabo G, Saha B: Alcohol’s Effect on Host Defense. Alcohol Res 2015, 37(2):159–170.

36. Trevejo-Nunez G, Kolls JK, de Wit M: Alcohol Use As a Risk Factor in Infections and Healing: A Clinician’s Perspective. Alcohol Res 2015, 37(2):177–184.

37. Pham CT: Neutrophil serine proteases: specific regulators of inflammation. Nat Rev Immunol 2006, 6(7):541–550.

38. Zamolodchikova TS, Tolpygo SM, Svirshchevskaya EV: Cathepsin G-Not Only Inflammation: The Immune Protease Can Regulate Normal Physiological Processes. Front Immunol 2020, 11:411.

39. Sabri A, Alcott SG, Elouardighi H, Pak E, Derian C, Andrade-Gordon P, Kinnally K, Steinberg SF: Neutrophil cathepsin G promotes detachment-induced cardiomyocyte apoptosis via a protease-activated receptor-independent mechanism. J Biol Chem 2003, 278(26):23944–23954.

40. Hoenderdos K, Condliffe A: The neutrophil in chronic obstructive pulmonary disease. Am J Respir Cell Mol Biol 2013, 48(5):531–539.

41. Vlodavsky I, Blich M, Li JP, Sanderson RD, Ilan N: Involvement of heparanase in atherosclerosis and other vessel wall pathologies. Matrix Biol 2013, 32(5):241–251.

42. Lerner I, Hermano E, Zcharia E, Rodkin D, Bulvik R, Doviner V, Rubinstein AM, Ishai-Michaeli R, Atzmon R, Sherman Y et al: Heparanase powers a chronic inflammatory circuit that promotes colitis-associated tumorigenesis in mice. J Clin Invest 2011, 121(5):1709–1721.

43. Planer D, Metzger S, Zcharia E, Wexler ID, Vlodavsky I, Chajek-Shaul T: Role of heparanase on hepatic uptake of intestinal derived lipoprotein and fatty streak formation in mice. PLoS One 2011, 6(4):e18370.

44. Vicente-Duenas C, Hauer J, Cobaleda C, Borkhardt A, Sanchez-Garcia I: Epigenetic Priming in Cancer Initiation. Trends Cancer 2018, 4(6):408–417.

45. Martucci C, Franchi S, Giannini E, Tian H, Melchiorri P, Negri L, Sacerdote P: Bv8, the amphibian homologue of the mammalian prokineticins, induces a proinflammatory phenotype of mouse macrophages. Br J Pharmacol 2006, 147(2):225–234.

46. Ring J: Fatal Effect of Eau Medicinale. Med Phys J 1811, 25(145):207.

47. Fung TT, McCullough ML, Newby P, Manson JE, Meigs JB, Rifai N, Willett WC, Hu FB: Diet-quality scores and plasma concentrations of markers of inflammation and endothelial dysfunction. The American Journal of Clinical Nutrition 2005, 82(1):163–173.

48. Chiuve SE, Fung TT, Rimm EB, Hu FB, McCullough ML, Wang M, Stampfer MJ, Willett WC: Alternative Dietary Indices Both Strongly Predict Risk of Chronic Disease. The Journal of Nutrition 2012, 142(6):1009–1018.

49. Huan T, Liu C, Joehanes R, Zhang X, Chen BH, Johnson AD, Yao C, Courchesne P, O’Donnell CJ, Munson PJ et al: A systematic heritability analysis of the human whole blood transcriptome. Hum Genet 2015, 134(3):343–358.

