## Supplemental Figures for "Blood transcriptomic biomarkers of alcohol consumption and cardiovascular disease risk factors: the Framingham Heart Study"

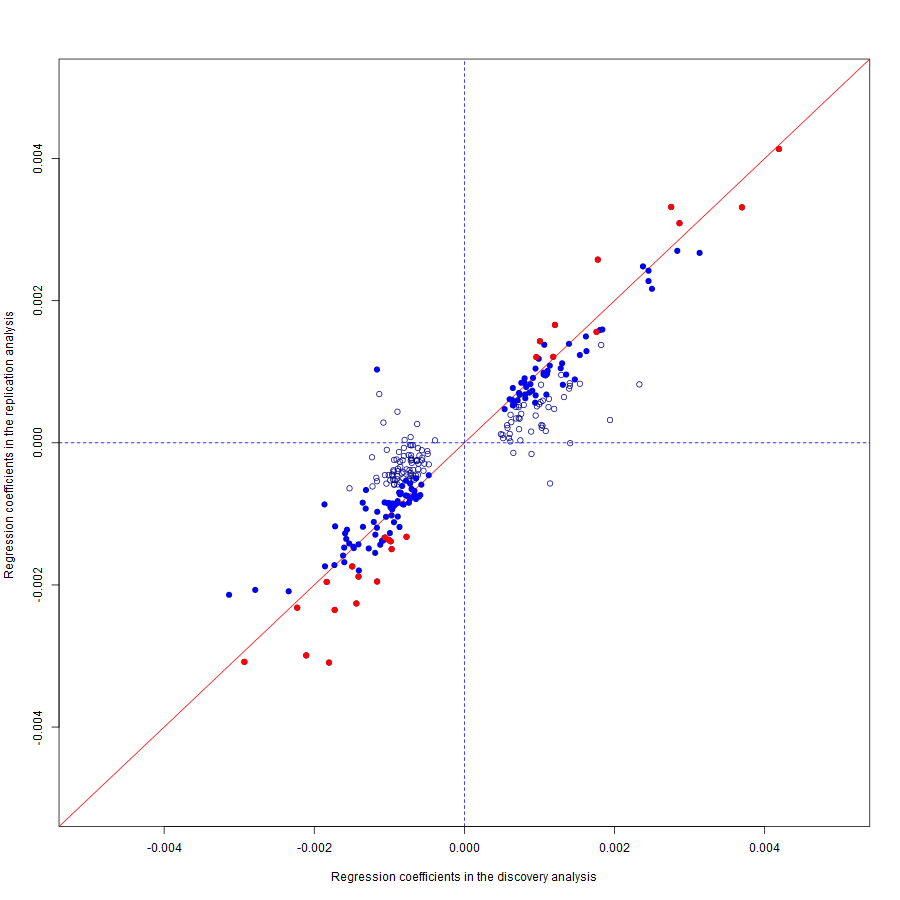

Regression coefficients in the discovery sample

Regression coefficients in the replication sample

**Supplemental Figure 1**. Comparison of regression coefficients of the 268 genes associated with alcohol consumption with FDR<0.05 in the discovery analyses. Blue colored dots are genes with *P* < 0.05 and red colored dots are significant genes (n = 25) after Bonferroni correction in the replication cohort.

Regression coefficients derived from analysis in all study samples (i.e., including heavy drinkers)

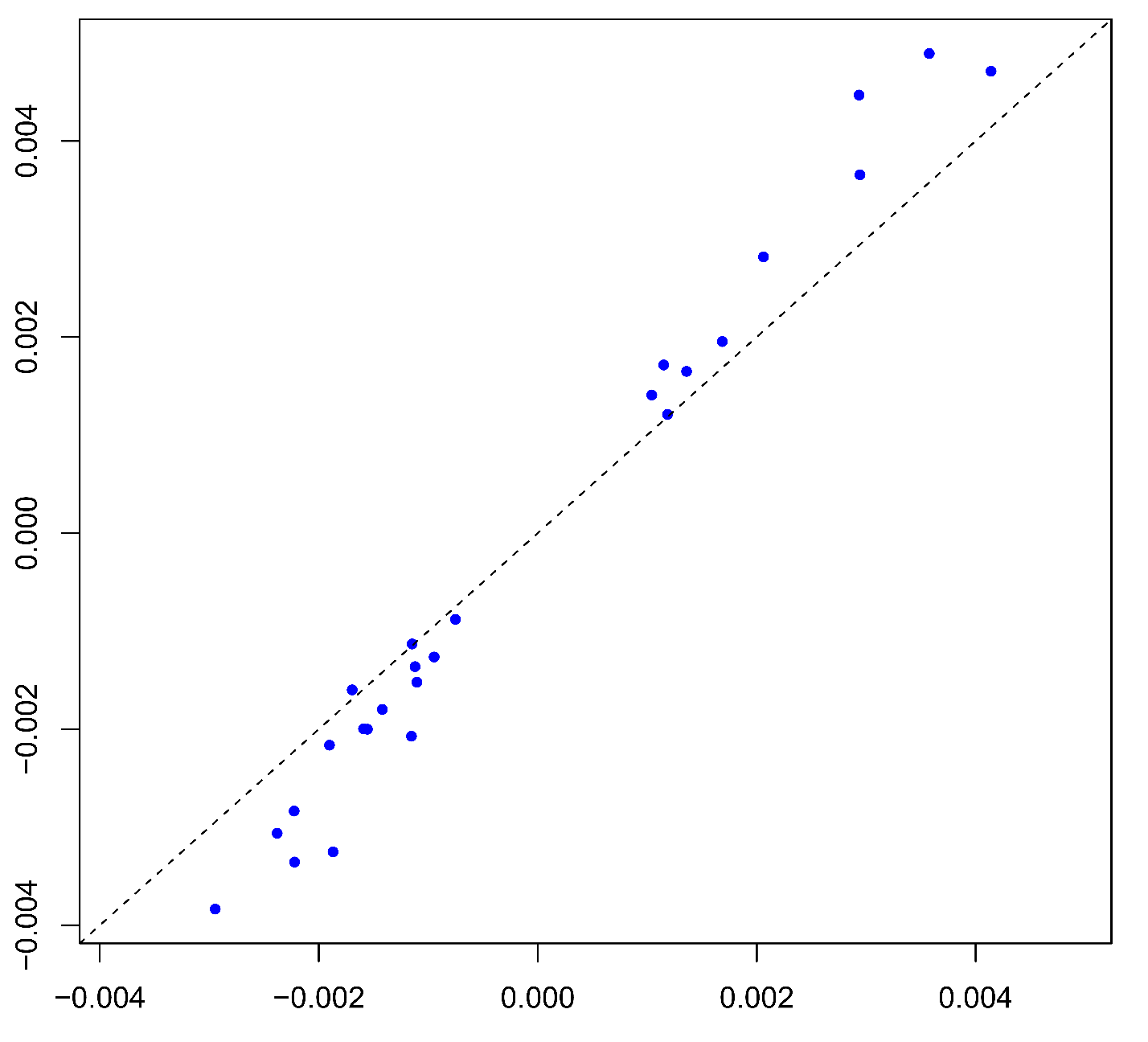

Regression coefficients derived from analysis in samples without heavy drinkers

Regression coefficients derived from analysis in all samples

**Supplemental Figure 2**. Comparison of regression coefficients of the 25 alcohol-associated genes in all study participants and in those after heavy drinkers were excluded.

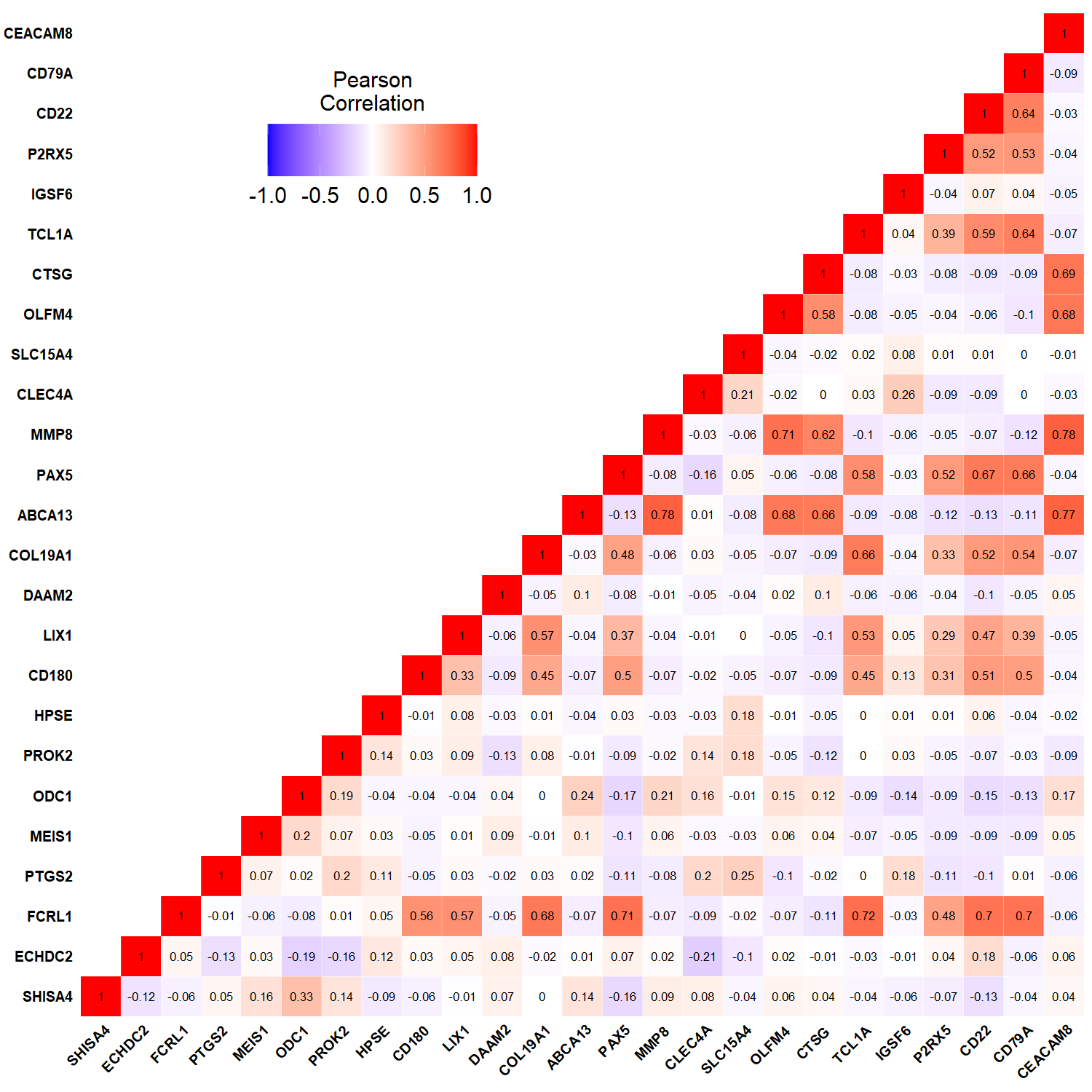

**Supplemental Figure 3**. Pairwise correlation coefficients of the 25 alcohol-associated genes

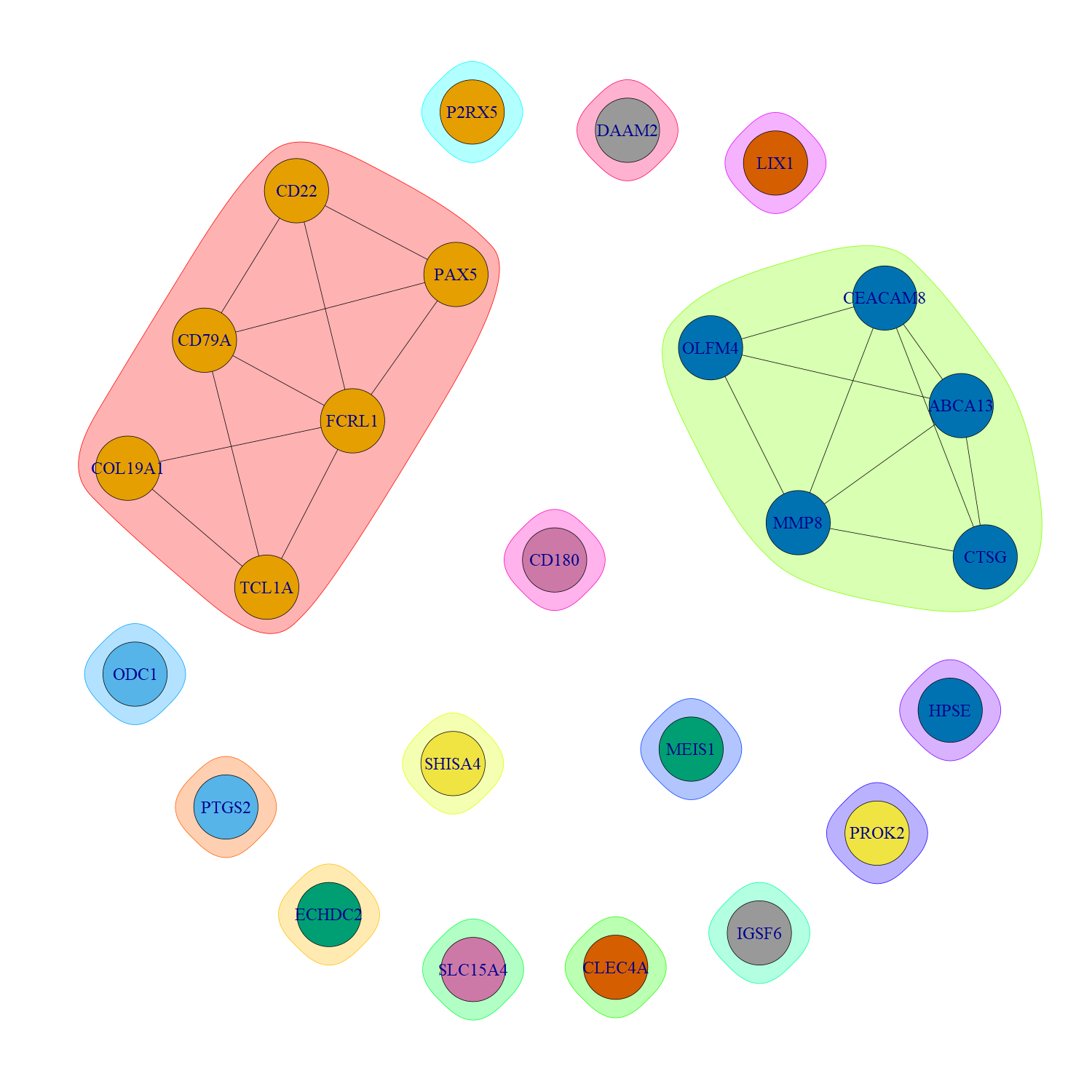

**Supplemental Figure 4**. Cluster visualization of the 25 alcohol-associated genes

Cross-sectional association: 1.17; 95%CI: 1.03, 1.34; *P* = 0.02

MR analysis: 0.11; 95%CI: 0.04, 0.18; P=0.002

Alcohol consumption

*CTSG*

BP/HTN

β = 0.0035

95%CI: 0.0027, 0.0044

P = 8.1e-16

HR = 1.14

95% CI: 1.04, 1.26

*P* = 0.007

Cross-sectional association: 2.14; 95%CI: 1.67, 2.76; *P* = 2.7e-9

MR analysis: 0.003; 95%CI: 0.002, 0.005; P=9.3e-5

Alcohol consumption

*HPSE*

BMI/Obesity

β = -0.0012

95%CI: -0.0016, -0.0007

P = 1.1e-7

HR = 0.80

95% CI: 0.71, 0.91

*P* = 4.6e-4

**Supplemental Figure 5**. Associations between alcohol consumption, expression levels of *CTSG* and *HPSE*, and CVD risk factors. Association between alcohol consumption and gene expression were from meta-analysis and association between alcohol consumption and CVD risk factors were reported in our previous study in FHS (PMID: 34159370)
